# Interest Holders-Driven Research Priorities in Sexual and Reproductive Health for Migrant and Refugee Adolescents and Young Adults in Australia

**DOI:** 10.1101/2025.07.11.25331354

**Authors:** Ahmed Shabbir Chaudhry, Patience Castleton, Tesfaye S. Mengistu, Negin Mirzaei Damabi, Afzal Mahmood, Rehana A Salam, Salima Meherali, Soumyadeep Bhaumik, Gizachew A. Tessema, Zelalem Mengesha, Humaira Maheen, Zachary Munn, Zohra S Lassi

## Abstract

**Background:** Despite sustained global commitment to sexual and reproductive health rights (SRHR), adolescents and young adults from migrant and refugee backgrounds continue to face disproportionate barriers to accessing appropriate services. In Australia’s multicultural landscape, systemic issues such as cultural stigma, limited sexual health literacy, and inadequate culturally responsive services exacerbate these disparities, highlighting the need for an interest-holder-centred and evidence-informed approach to sexual and reproductive health (SRH) research prioritisation.

**Methods:** This study employed the James Lind Alliance (JLA) Priority Setting Partnership (PSP) methodology to collaboratively identify and prioritise key research uncertainties in SRH for migrant and refugee adolescents and young adults in Australia. Guided by a multidisciplinary steering committee, the process encompassed five structured stages: partnership formation, literature-based identification of uncertainties, interest holders’ consultation and refinement, an interim prioritisation survey, and a final consensus workshop. A rapid literature review generated 83 research questions across five domains and twelve thematic areas, which were assessed via two rounds of structured online surveys. Participants included youth interest holders (adolescents and young adults aged 18–24 from migrant and refugee backgrounds), alongside diverse professional interest holders from healthcare, academic, governmental, and community sectors.

**Results:** A total of 92 participants (31 youth and 61 professionals) completed the initial survey, generating 83 research uncertainties that were refined into 18 indicative questions across five SRH domains. Among youth interest-holders, 74.2% identified as female and 25.8% as male. Professional interest-holders were more gender-diverse, with 49.2% female, 32.8% male, 8.2% transgender, and 9.8% preferring not to disclose. Youth participants predominantly identified psychosocial, community-based, and education-related concerns, while professionals drawn from academic/research (37.7%), health industry organizations (13.1%), healthcare services (44.3%), and NGOs emphasised system-level barriers and challenges in service access. Using median scores, eight questions emerged as shared high priorities across both groups. In the final consensus phase, 11 questions were identified as top priorities, with three receiving high-priority ratings from both youth and professionals. Key areas included male SRH, psychosocial contributors to gender-based violence, culturally tailored education, and community-based maternal health support. Notably, youth participants consistently rated more items as high priority than professionals, reflecting divergent perspectives shaped by lived experience. These results provide an empirically grounded, interest holders-informed SRH research agenda for migrant and refugee youth in Australia.

**Conclusion:** This interest-holders driven priority setting initiative highlights the pressing need for tailored SRH research responsive to the lived experiences of adolescent and young adults of migrant and refugee background in Australia. The identified priorities serve as a foundation for future evidence-based interventions and policy development to advance health equity and service inclusivity.

## INTRODUCTION

Sexual and Reproductive Health and Rights (SRHR) encompass the right of every individual to make informed choices about their body, sexuality, and reproduction free from discrimination, coercion, or violence and are universally recognised as fundamental to personal wellbeing, public health, and sustainable development (1). The concept of SRHR was formally articulated in the Programme of Action at the International Conference on Population and Development (ICPD) in 1994, SRHRs have since been reaffirmed as fundamental human right (2). However, despite decades of global advocacy, policy efforts, and international commitments, progress toward equitable access to comprehensive SRH services remains uneven due to structural inequalities, sociopolitical resistance, and systemic underinvestment (1), particularly for adolescents, young people, and marginalised groups such as migrants and refugees. Migrant communities, in particular, experience disproportionate and persistent obstacles in accessing sexual and reproductive health (SRH) services due to various pervasive challenges like socioeconomic and language barriers, fear of deportation, discrimination, and absence of culturally responsive care (3).

Adolescents are the most vulnerable group as they transition through significant physical, emotional, and social changes with lasting implications for health (4). Many young people face increased SRHR vulnerabilities due to gender-based violence, early and forced marriage, unintended pregnancy, and limited access to essential services (1). Nearly 29% of girls aged 15-19 who are currently or previously in intimate relationships have experienced intimate partner violence. Although violence against women is deeply rooted in society, effective responses, especially preventive measures, have not been implemented widely (1). In many low-resource contexts, child marriage contributes to early motherhood and heightened susceptibility to Human Immunodeficiency Virus (HIV) and sexually transmitted infections (STIs). Yet, despite the increasing need, over half of adolescent girls in developing regions who wish to avoid pregnancy are unable to access modern contraception primarily due to legal restrictions, stigma, and provider bias. These structural and systemic barriers to contraception access reflect a broader pattern of constrained SRH rights worldwide (1, 5-7). Importantly, access to abortion is increasingly restricted not only in low-resource contexts but also in high-income countries, disproportionately affecting young, poor, and marginalized populations, and mirroring global patterns of SRH inequity (8). Even in legally permissive contexts like Australia, women particularly those from rural, migrant, and marginalized communities face significant non-legal barriers to abortion access, including high out-of-pocket costs, geographic inaccessibility, and practitioner-related obstacles, all of which continue to undermine reproductive autonomy despite legislative reforms (9). These intersecting barriers compromise adolescents’ autonomy, health outcomes, and life trajectories.

SRH vulnerabilities are further exacerbated by global patterns of migration and displacement, which intensify the exposure to instability, disrupted care, and rights violations (3). Migration now defines the global landscape, with 3.6% of the world’s population, approximately 281 million individuals, living outside their country of origin (10). Among these, 117.3 million have been forcibly displaced, including 43.4 million refugees (11). Australia has one of the highest proportions of overseas-born residents among high-income countries. As of June 2024, 31.5% of Australia’s population, equivalent to 8.6 million people, were born overseas (12). This marks the highest proportion since 1893 and underscores Australia’s sustained reliance on international migration to offset low natural population growth, fill critical labour shortages particularly in sectors like healthcare and technology and attract skilled individuals who contribute significantly to economic productivity and regional development (12, 13). Such demographic diversity underscores the need for inclusive, culturally responsive health policies, particularly in addressing SRHR for migrant and refugee populations.

Within this demographic context, growing empirical evidence underscores the persistent systemic and sociocultural barriers that compromise equity of SRHR for migrant and refugee youth (14). A recent systematic review of Australian literature identified a constellation of interrelated challenges, including limited SRHR knowledge, cultural and religious stigma, structural impediments to service access, and pervasive mistrust of healthcare systems (15). These barriers are further exacerbated by experiences of discrimination, social exclusion, and a widespread lack of culturally safe care within clinical environments. The review emphasises that these multi-level barriers intersect across individual, interpersonal, institutional, and societal domains, collectively shaping the SRHR experiences, choices, and outcomes of this population (15).

In response to these entrenched challenges, there is increasing recognition of the imperative to meaningfully involve patients and the public across all stages of the healthcare and research continuum (16). This shift towards participatory approaches positions individuals with lived experience not merely as recipients of care, but as co-producers of knowledge whose insights are critical to shaping relevant, context-sensitive research agendas (17). When combined with the expertise of service providers, this collaborative model ensures that interventions are both grounded in real-world experience and aligned with systemic capacities (18). Guided by this framework, the present study sought to identify and prioritise SRH research priorities for adolescent and young adults from migrant and refugee backgrounds in Australia, drawing on diverse interest-holder perspectives, including young people themselves, as well as community members, service providers, researchers, and policy influencers.

## METHOD

This study was conducted as a Priority Setting Partnership (PSP) following the standardised methodology developed by the James Lind Alliance (JLA) (19). PSP aims to bring together patients, carers and clinicians to identify and prioritise unanswered questions for future research(19). This inclusive and systematic approach identifies and addresses evidential uncertainties, which are defined as questions about the effects of treatments or interventions for which reliable, up-to-date systematic reviews are lacking or where existing evidence is conflicting or inconclusive, across a broad range of health domains (19). The JLA approach consists of the following key stages: 1) project initialisation and partnership formation, 2) collection of uncertainties, 3) data processing and verification of uncertainties through consultation, 4) interim prioritisation through interest holders’ survey, and 5) final consensus to establish definitive priorities (19). Each stage adheres to established JLA protocols to ensure methodological rigour and transparency throughout the priority-setting process.

While there is no formal reporting guideline specific to JLA PSPs, this study was reported using the Reporting guideline for priority setting of health research (REPRISE) to enhance reporting clarity and transparency **(Supplementary file 1)** (20). In this study, we adopt the term “interest-holders” rather than the more commonly used “stakeholders” to emphasise the normative legitimacy of those involved in, or affected by, the health issue under investigation (21). This terminology shift is intended to address concerns that the term “stakeholder” may obscure important power dynamics and imply an equivalence of influence among groups that often differ vastly in their ability to shape health research and policy. Interest-holders are defined as “individuals or groups who have a legitimate interest in a health issue either because they are directly affected by it or because they hold decision-making responsibilities that could be informed by research evidence” (21). This reframing encourages more inclusive and equitable participation by recognising those with lived experience and moral or professional accountability, rather than privileging institutional authority or policy influence alone (21). In the context of this PSP, interest-holders include both adolescents and young adults from migrant and refugee backgrounds, who are directly impacted by such decisions, as well as healthcare professionals, researchers, policymakers, and organisational, community caregivers and leaders, who play key roles in the development and implementation of SRH services and policies. For clarity, we refer to adolescents and young adults as “youth interest-holders” and researchers, policymakers, and organisational leaders as “professional interest-holders” throughout this paper.

The priority-setting process was implemented across the following integrated methodological steps, described in detail below:

### 1. Project initialisation and partnership formulation

A steering committee was established with diverse expertise comprising researchers specialised in SRH, adolescent and youth health, and migrant and refugee health; healthcare providers with relevant clinical experience; and young adult representatives from the target population.

### 2. Collection of uncertainties

To identify the top priorities of adolescents and young adults (aged 18 to 24 years) from migrant and refugee backgrounds in SRH, a rapid literature review employing a systematic approach was conducted to balance methodological rigour with practical constraints of time and resources, enabling efficient synthesis of existing evidence while upholding transparency and reproducibility (22). Our review examined research published in English between 2000 and 2024, including empirical studies addressing SRH within this specific population in Australia. Studies unrelated to SRH or those not reporting disaggregated data for migrant and refugee youth were excluded. Searches were conducted across three academic databases, namely PubMed, CINAHL, and Web of Science, to minimise publication bias and ensure comprehensive literature coverage. Screening was conducted by two reviewers (PC and TM) based on predefined inclusion and exclusion criteria, with discrepancies resolved through discussion or consultation with a senior author (ZSL).

### 3. Data Processing and verification of uncertainties through consultation

We systematically identified research gaps within the literature, which were then presented to the steering group for collaborative analysis. Through structured discussion, the steering group categorically refined and consolidated these uncertainties into 83 research questions (**see Supplementary file 2**: for details on the distinction between research gaps and uncertainties). This analytical process resulted in 12 distinct thematic areas, which were further consolidated into five overarching domains encompassing various aspects of SRH among adolescents and young adults from migrant and refugee backgrounds (**Box 1**). These domains capture the nature and focus of each question, whether related to service access, interventions, health literacy, or vulnerabilities.

#### Box 1

**SRH Thematic Areas and Consolidated Domains**

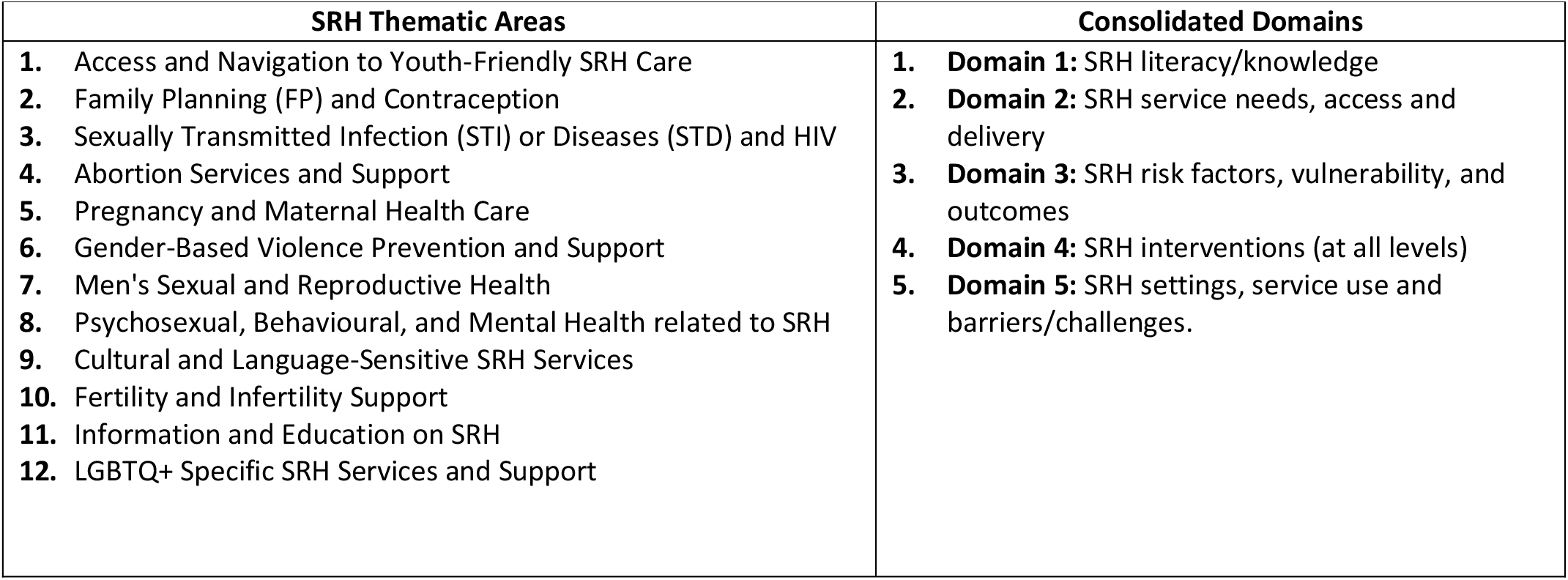

### 4. Interim prioritisation through the Interest-holders survey (Initial survey)

The first phase of data collection involved an online survey conducted between 23 Sep 2024 and 24 Jan 2025, aimed at capturing perspectives from a diverse and representative sample of both youth and professional interest-holders.

The survey comprised two main components. The first was a demographic questionnaire that gathered information on participants’ age range, gender, sector of affiliation, and years of experience within that sector. The second component was a research prioritisation questionnaire, which presented participants with 83 evidence uncertainties structured across five key domains of SRH, reflecting the scope of the priority-setting exercise (**see Supplementary File 1**).

Participants were asked to rate each of the 83 research questions using a 9-point Likert scale, with 1–3 indicating low priority, 4–6 moderate priority, and 7–9 high priority. This rating approach followed the standard methodology of the JLA for assessing the perceived importance of health research questions (19). Responses were analysed to identify which questions were viewed as most critical, drawing on both experiential and professional perspectives.

#### Participants and Recruitment of Interest-holders

Following the conceptual framework of this study, participants were categorised into two distinct groups of interest-holders: youth interest-holders and professional interest-holders.

- **Youth interest-holders:** Adolescents and young adults aged 18 to 24 years from migrant and refugee communities living in Australia were included as youth interest-holders to ensure that those directly affected by SRH policies and services could meaningfully contribute to the priority-setting process. Recruitment was facilitated through partnerships with community-based and cultural organisations, schools, and healthcare providers serving diverse populations. These settings were selected to foster trust and ensure the inclusion of youth who are often excluded from formal research and policy processes. Their involvement aimed to capture the intersectional barriers they face in accessing SRH care, including cultural norms, language barriers, limited health literacy, and systemic discrimination.
- **Professional interest-holders:** Individuals engaged in research, policy, service delivery, or advocacy related to SRH among migrant and refugee populations. Participants were recruited from a broad range of sectors, including academic institutions, public and private healthcare services, international and national non-governmental organisations, and government agencies such as the Australian Department of Health and SA Health. Community-based SRH providers, such as Shine SA, were also included. Recruitment was conducted through web-based searches, referrals from the project’s steering committee, and direct outreach to relevant organisations. Their inclusion ensured the study captured diverse perspectives from those involved in shaping, delivering, and evaluating SRH interventions and policies at both local and national levels.

To maximise participation, the survey was distributed through a combination of direct email invitations (to over 200 participants all over Australia) and targeted promotion on social media platforms, including Instagram, Facebook, and X (formerly Twitter). Additionally, the survey was circulated through existing networks, including refugee and migrant centres, youth organisations, and academic institutions. Recruitment of youth interest-holders was supported through collaboration with migrant and refugee organisations, schools and cultural or community hubs. Community and religious leaders played an instrumental role in identifying eligible participants and promoting the survey among migrant and refugee youth. Upon confirmation of eligibility, all participants were invited to complete the survey online. Demographic data were monitored throughout the collection period to identify gaps in representation and guide further outreach efforts to underrepresented groups.

Prior to the official launch, the survey was pilot-tested with members of the project’s steering committee and a small group of 12 or 15 individuals from each group of youth and professional interest-holders. Participants were recruited through convenience sampling and provided feedback on the clarity, accessibility, and cultural appropriateness of the survey content. Based on their input, minor revisions were made to ensure the survey was clear, acceptable, and appropriate for the intended population.

### Data Analysis: Refinement and Interim Prioritisation

The data analysis process for the initial survey round involved several distinct steps to refine, assess, and prioritise interim research uncertainties identified by interest-holders. Following the initial survey, responses were analysed using median scores to determine the perceived importance of each of the 83 research questions. To account for the perspectives of both key groups, median scores were calculated separately for youth interest-holders and professional interest-holders. This stratified approach allowed the research team to identify interim high-priority uncertainties within and across participant categories.

Research questions with consistently high median scores were identified as top uncertainties. These questions were reviewed by the study’s steering committee to ensure they reflected the priorities and experiences of participants and aligned with the broader objectives of the PSP.

### 5. Final consensus to establish definitive priorities (second survey)

The refined interim priorities were used to develop a final prioritisation survey aimed at validating and ranking the most critical research questions. Participants were provided with the top-ranked priorities from the initial survey and asked to re-rank them based on perceived urgency and importance. To support informed decision-making, a brief summary of these priorities was provided at the beginning of the survey. This round of the survey was conducted between 05 Feb 2025 and 11 Mar 2025.

The final survey was distributed using the same multi-platform recruitment strategy employed in the initial round, targeting both youth and professional interest-holders. Demographic data were again collected to monitor the composition of respondents and enable subgroup analysis.

Survey responses were analysed using a similar approach as in the initial round to determine the overall rankings of each question. Results were stratified by participant group to ensure equitable representation of both youth and professional interest-holders’ perspectives. The ranked lists were then reviewed by the steering committee, which confirmed the final set of questions to be advanced to the consensus workshop.

The final phase of data analysis focused on reaching consensus on the definitive set of research priorities. This was conducted through online consultation with a steering committee and individuals from each youth and professional interest-holders backgrounds, where members were invited to review the rankings, provide feedback, and raise any concerns or suggestions. Following this iterative review and the incorporation of relevant input, a final consensus was reached. This concluding step ensured that the prioritised list reflected broad agreement and equitable contributions from both youth and professional interest-holders.

### Ethical Consideration

Formal ethics approval was obtained from the University of Adelaide Human Research Ethics Committee (H-2023-234) before commencing project activities.

## RESULTS

### Results

A total of 92 participants responded to the initial survey, representing diverse demographics and organisational types (**Table 1**).

**Table 1:** Characteristics of youth and professional Interest-Holders (N = 92)

| Characteristic | Youth Interest Holders<br>(n=31) | Professional Interest Holders<br>(n=61) |
| --- | --- | --- |
| **Gender Identity** |  |  |
| Female | 23 (74.2%) | 30 (49.2%) |
| Male | 8 (25.8%) | 20 (32.8%) |
| Transgender | 0 | 5 (8.2%) |
| Prefer not to say | 0 | 6 (9.8%) |
| **WHO Region (Country of Origin)** |  |  |
| South-East Asia Region | 13 (41.9%) | 23 (37.7%) |
| Eastern Mediterranean Region | 8 (25.8%) | 13 (21.3%) |
| African Region | 4 (12.9%) | 2 (3.2%) |
| Region of the Americas | 3 (9.6%) | 11 (18.2%) |
| European Region | 2 (6.5%) | 4 (6.5%) |
| Western Pacific Region | 1 (3.2%) | 8 (13.11%) |
| **Industry Affiliation (professionals only)** |  |  |
| Academic/Research |  | 23 (37.7%) |
| Health Industries (e.g., SHINE SA) |  | 8 (13.1%) |
| Healthcare Services (e.g., clinics) |  | 27 (44.3%) |
| International NGO<sup>^</sup> |  | 2 (3.3%) |
| National NGO<sup>^</sup> |  | 1 (1.6%) |

<sup>^</sup>Non-Governmental Organisation (NGO)

### Initial Survey Results

The initial survey results highlighted 18 key research priorities across five SRH domains (**Table 2**). Among these, 16 high-priority questions were identified by professional interest holders and 10 by youth interest holders. Together, eight common questions were ranked as high priority by both interest holders. Participants were asked to rate each question based on perceived importance, with median scores and inter quartile ranges (IQRs) used to determine relative priority and consensus levels, providing a dual indication of importance and internal agreement.

**Table 2:**
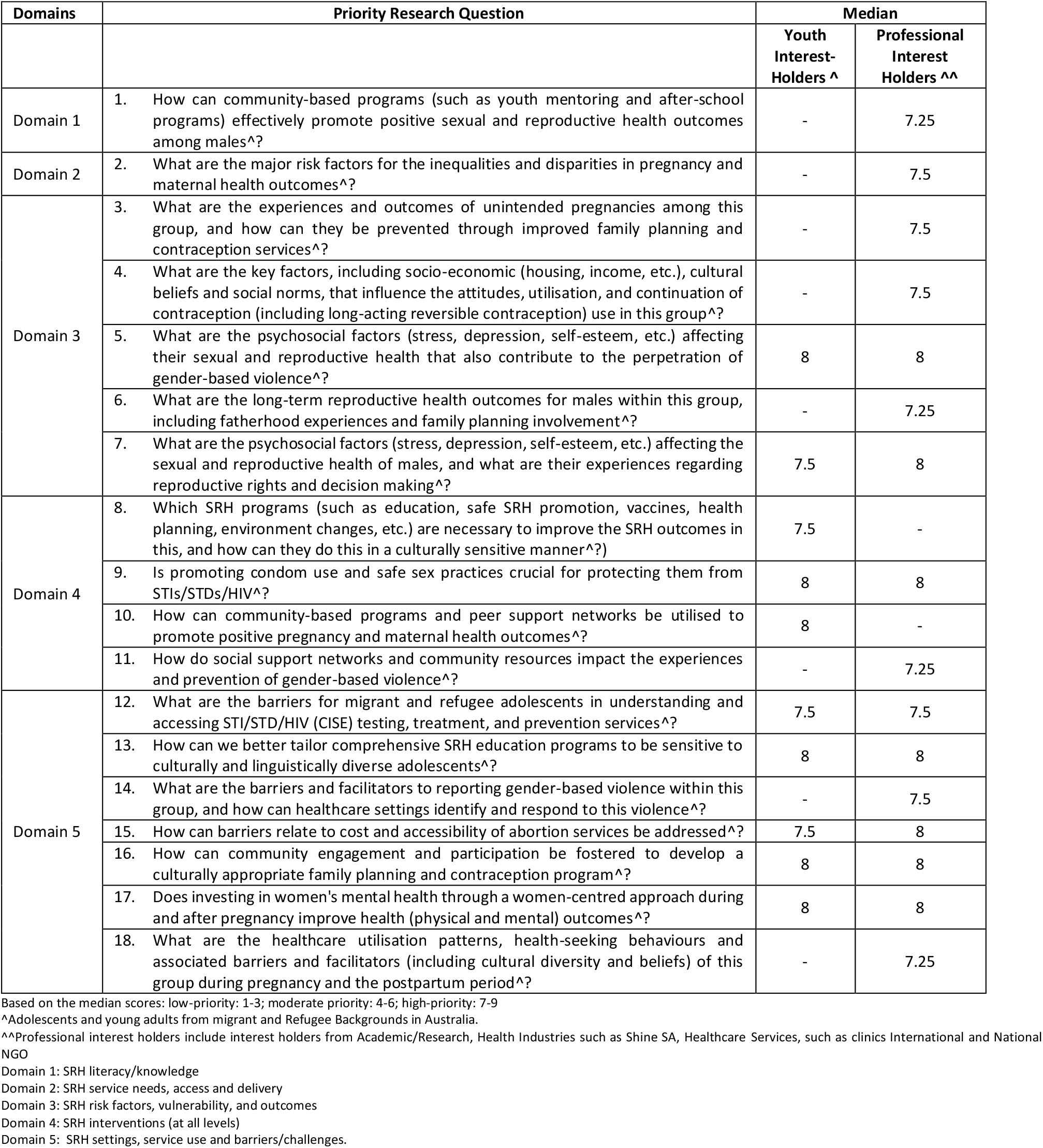
Top Research Priorities from the first round of survey on SRH for adolescents and young adults from Migrant and Refugee Backgrounds in Australia.

### Final Survey Results

The final prioritisation phase involved 92 interest-holder responses. While Domain 2 (SRH service needs, access and delivery) did not yield any questions rated as high priority during the consensus stage, eleven questions across Domains 1 to 5, excluding 2, were confirmed as high-priority items (**Table 3**).

**Table 3:**
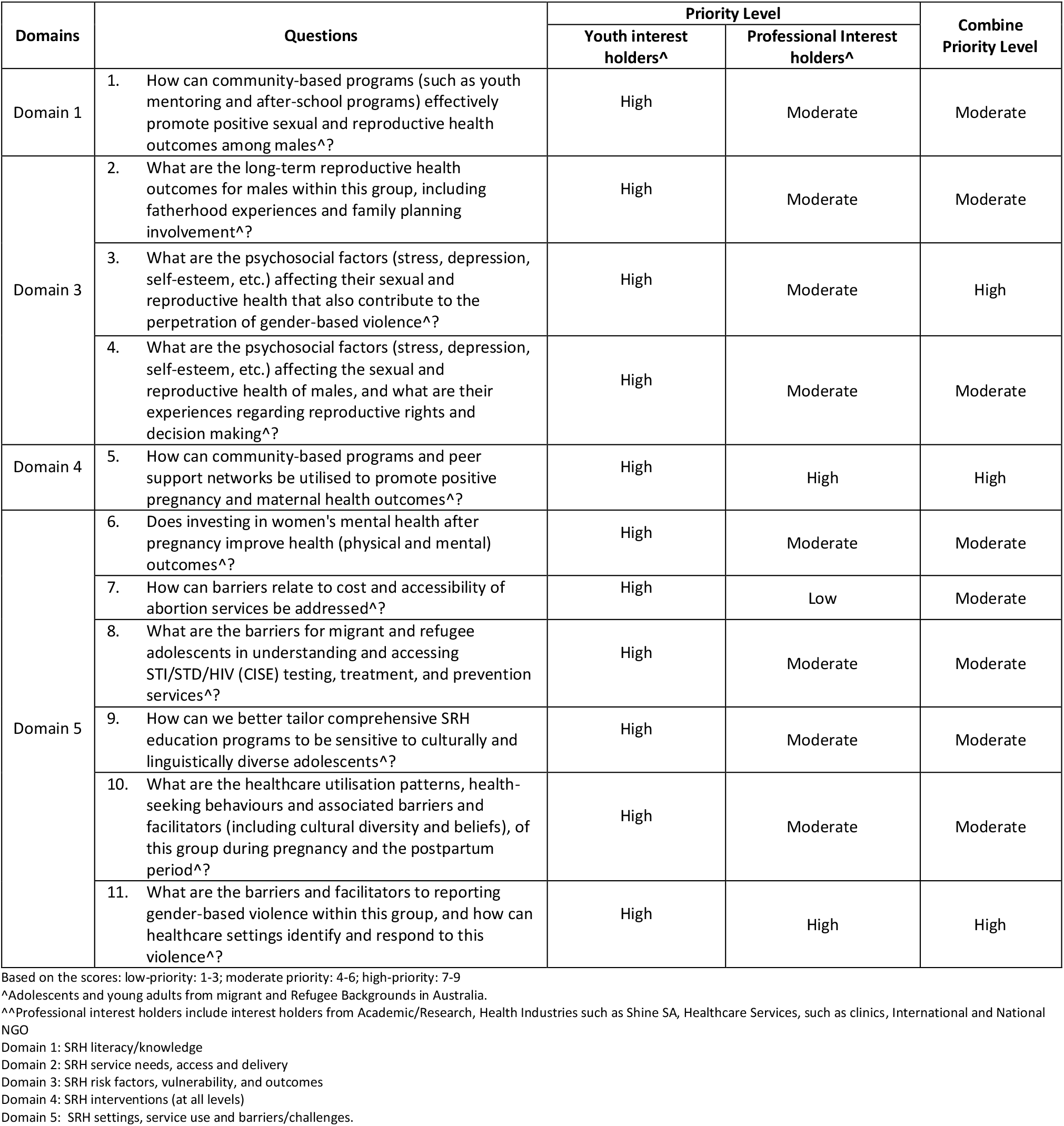
Final top research priorities on SRH for adolescents and young adults from migrant and Refugee Backgrounds in Australia.

| Domains | Questions | Priority Level |  | Combine Priority Level |
| --- | --- | --- | --- | --- |
|  |  | Youth interest holders^ | Professional Interest holders^ |  |
| Domain 1 | 1. How can community-based programs (such as youth mentoring and after-school programs) effectively promote positive sexual and reproductive health outcomes among males^? | High | Moderate | Moderate |
| Domain 3 | 2. What are the long-term reproductive health outcomes for males within this group, including fatherhood experiences and family planning involvement^? | High | Moderate | Moderate |
|  | 3. What are the psychosocial factors (stress, depression, self-esteem, etc.) affecting their sexual and reproductive health that also contribute to the perpetration of gender-based violence^? | High | Moderate | High |
|  | 4. What are the psychosocial factors (stress, depression, self-esteem, etc.) affecting the sexual and reproductive health of males, and what are their experiences regarding reproductive rights and decision making^? | High | Moderate | Moderate |
| Domain 4 | 5. How can community-based programs and peer support networks be utilised to promote positive pregnancy and maternal health outcomes^? | High | High | High |
| Domain 5 | 6. Does investing in women's mental health after pregnancy improve health (physical and mental) outcomes^? | High | Moderate | Moderate |
|  | 7. How can barriers relate to cost and accessibility of abortion services be addressed^? | High | Low | Moderate |
|  | 8. What are the barriers for migrant and refugee adolescents in understanding and accessing STI/STD/HIV (CISE) testing, treatment, and prevention services^? | High | Moderate | Moderate |
|  | 9. How can we better tailor comprehensive SRH education programs to be sensitive to culturally and linguistically diverse adolescents^? | High | Moderate | Moderate |
|  | 10. What are the healthcare utilisation patterns, health-seeking behaviours and associated barriers and facilitators (including cultural diversity and beliefs), of this group during pregnancy and the postpartum period^? | High | Moderate | Moderate |
|  | 11. What are the barriers and facilitators to reporting gender-based violence within this group, and how can healthcare settings identify and respond to this violence^? | High | High | High |

Based on the scores: low-priority: 1-3; moderate priority: 4-6; high-priority: 7-9
^^Professional interest holders include interest holders from Academic/Research, Health Industries such as Shine SA, Healthcare Services, such as clinics, International and National NGO
Domain 1: SRH literacy/knowledge
Domain 2: SRH service needs, access and delivery
Domain 3: SRH risk factors, vulnerability, and outcomes
Domain 4: SRH interventions (at all levels)
Domain 5: SRH settings, service use and barriers/challenges.

Among these, two questions were rated as high priority by both youth and professional interest-holders, reflecting strong consensus across experiential and professional perspectives. These were:

1. “How can community-based programmes and peer support networks be utilised to promote positive pregnancy and maternal health outcomes?” (Domain 4), and
2. “What are the barriers and facilitators to reporting gender-based violence within this group, and how can healthcare settings identify and respond to this violence?” (Domain 5).

Conversely, only one question, “How can barriers related to cost and accessibility of abortion services be addressed?” (Domain 5), was rated high by youth interest-holders but low by professional participants, resulting in a combined priority level of moderate. A notable pattern emerged in the aggregate prioritisation levels: while the majority of the questions attained a combined priority level of *moderate*, only three questions were classified as *high priority* in the final synthesis. In addition to the two items achieving dual-group consensus, a third question attained a combined high priority status despite only moderate professional interest: “What are the psychosocial factors (stress, depression, self-esteem, etc.) affecting their sexual and reproductive health that also contribute to the perpetration of gender-based violence?” (Domain 3).

## DISCUSSION

The results identified key research priorities in SRH for migrant and refugee adolescents and young adults in Australia through a multi-stage, participatory process. Initial data collection yielded 83 research uncertainties, which were refined into 18 interim indicative questions across five domains. Of these, eight were identified as high priorities by both youth and professional interest-holders. The final prioritisation phase, further refined these uncertainties, culminating in 11 final priority questions established through consensus. These questions constitute an interest-holder-centred and evidence-informed research agenda that responds directly to the lived experiences and systemic realities of migrant and refugee youth in Australia. This methodological approach enhances the alignment of future research with population-level needs and provides a robust foundation for policymaking, clinical practice, and resource allocation.

A salient pattern emerging from the findings is the consistent tendency of youth interest-holders to assign higher priority ratings across a broad spectrum of SRH topics compared to professionals’ interest-holders. This divergence underscores fundamental differences in the perception and evaluation of health needs between the two groups. Professional interest-holders, often operating within institutional and policy frameworks, may prioritise issues based on structural feasibility and resource allocation considerations. In contrast, youth participants, drawing from their lived experiences, are more likely to prioritise issues that are immediately visible or acutely felt in their everyday lives. This divergence in prioritisation may also be influenced by varying levels of SRH literacy among youth. Evidence suggests that limited health literacy can affect individuals’ ability to accurately assess health priorities, potentially leading to an emphasis on more familiar or surface-level concerns over systemic determinants (23). For instance, a study conducted among adolescents found that over 74% had limited SRH literacy, impacting their understanding and prioritisation of SRH issues (24).

In the following sections, the prioritised questions are discussed thematically rather than by domain to better reflect conceptual linkages and shared contextual features across issues to present a more coherent, experience-aligned interpretation of the identified priorities.

### Sexual and Reproductive Health Literacy among Migrant and Refugee Youth

SRH literacy, classified under Domain 1, was not ranked as a high priority by either youth or professional interest-holders in the final consensus. This omission is particularly striking in light of the pervasive evidence documenting poor SRH literacy among culturally and linguistically diverse populations in Australia (25). It raises critical questions: do participants assume that literacy-based interventions are already in place, or does low literacy itself restrict awareness of how fundamental such knowledge is to achieve SRH rights and outcomes? This suggests that health research priorities are shaped not exclusively by objective need but also by participants’ perception, awareness, internalised cultural norms and values, and the visibility of issues within their cognitive frameworks.

Nonetheless, it is significant that one literacy-related question—*”How can we better tailor comprehensive SRH education programs to be sensitive to culturally and linguistically diverse adolescents?”* was prioritised under Domain 5. This reflects an implicit concern with literacy as it intersects with service delivery and cultural responsiveness. Rather than viewing SRH literacy as an isolated educational gap, participants appeared to value it in applied, relational contexts, particularly where communication, trust, and cultural appropriateness are critical. This underscores the importance of recognising health literacy as a contextualised and embodied experience, and not merely a cognitive or informational deficit.

### Male-Specific Sexual and Reproductive Health Priorities

The question, *“What are the long-term reproductive health outcomes for adolescent and young males, including fatherhood experiences and family planning involvement?”* reflects a critical gap in our understanding of the male continuum of reproductive health, particularly in migrant and refugee contexts. As highlighted in a scoping review on the SRH of migrant and refugee men in Australia, existing research is disproportionately focused on HIV and STIs, with limited attention to broader reproductive health domains such as fertility, contraception, and men’s roles in parenting and family planning (26). This narrow focus overlooks important aspects such as fatherhood experiences, shared decision-making in contraception, and psychosocial aspects of male reproductive roles.

International agencies such as the Partnership for Maternal, Newborn, and Child Health (PMNCH) (27), the World Health organisation (WHO) (28), and the Breakthrough ACTION advocacy tool (2019) (29) have consistently underscored the limited involvement of men in reproductive health, particularly during antenatal and parenting phases. These reports highlight that male engagement remains low due to entrenched cultural norms, health system gaps, and a lack of tailored programming. They advocate for intentionally integrating men as both users and partners in reproductive health, not only to improve male outcomes but also to enhance overall family and community wellbeing.

In this context, the prioritisation of the question *“How can community-based programs (such as youth mentoring and after-school programs) effectively promote positive sexual and reproductive health outcomes among males?”* highlights a promising avenue for addressing these longstanding service gaps. Community-based initiatives offer an alternative to formal health systems, creating culturally safe, trust-based platforms for engagement, especially critical for young men from backgrounds where SRH conversations are highly stigmatised. This priority also echoes the recommendations from a recent scoping review, which points to a lack of accessible, male-specific education and support structures in SRH services for migrant and refugee populations (26). By centring male voices and lived experiences in informal, community-embedded settings, such programs can help bridge the disconnect between service provision and the everyday realities of marginalised male youth.

The inquiry into *“What are the psychosocial factors (stress, depression, self-esteem, etc*.*) affecting the sexual and reproductive health of males and what are their experiences regarding reproductive rights and decision making?”* highlights the intersection of mental health and SRH. This relationship is well-documented in a 2023 scoping review, which found that mental health and psychosocial wellbeing are inextricably linked to SRH outcomes among adolescents and young people (30). The review emphasises that psychosocial challenges, such as emotional distress, depression, anxiety, stress symptoms, and low self-worth, are prevalent and significantly affect SRH. It advocates for integrated interventions that address these psychological challenges within SRH service delivery, especially in settings burdened by stigma, discrimination, and systemic barriers to care. Such approaches are crucial for improving both mental health and SRH outcomes in this demographic.

### Sexual and Reproductive Health Priorities

The prioritisation of the question, *“How can community-based programmes and peer support networks be utilised to promote positive pregnancy and maternal health outcomes?”* underscores the recognised need for culturally sensitive and accessible interventions tailored to migrant and refugee populations. Community-based models, such as the Refugee Midwifery Group Practice in Australia, have demonstrated improvements in maternal and neonatal outcomes by providing continuity of care and culturally appropriate services (31). These programmes often leverage peer support mechanisms, which have been effective in enhancing engagement and trust among migrant communities. Additionally, initiatives like the Cross-Cultural Workers (CCWs) in Maternity and Child and Family Health Services have been associated with positive experiences and high satisfaction rates among migrant and refugee women, indicating the potential of such models to improve accessibility and responsiveness of maternity care (32).

The question, *“How can barriers related to cost and accessibility of abortion services be addressed?”* draws attention to entrenched inequities in reproductive healthcare access, particularly for migrant and refugee populations. Despite progressive legal reforms across Australian jurisdictions, substantial disparities persist. Migrant and refugee women face considerable obstacles, including high out-of-pocket costs, inconsistent availability of abortion services, particularly in public hospitals and geographic inequities, with limited-service provision in regional and rural areas(9). These systemic factors significantly constrain equitable access(9).

This prioritisation reveals a noteworthy divergence in how different groups perceive SRH needs. While youth interest-holders ranked abortion access and cost as a high priority, professional participants did not, resulting in a moderate combined score. This discrepancy raises important questions about how structural issues, such as financial and geographic inaccessibility, facing marginalised populations are understood and valued by professionals. It invites critical reflection on whether institutional actors may inadvertently deprioritise issues that are deeply felt by service users but less visible within system-level planning. These differences are instructive. The abortion access question exemplifies concerns that strongly resonate with youth from marginalised backgrounds, even when professionals place less emphasis on them. This misalignment suggests a potential blind spot in professional frameworks, whereby pressing social or economic barriers may be overlooked in favour of priorities more closely aligned with institutional perspectives. Recognising and addressing these gaps is essential to ensure that SRH policies and research agendas remain firmly grounded in the lived realities of those most affected.

The prioritisation of the question, *“What are the barriers for migrant and refugee adolescents in understanding and accessing STI/STD/HIV testing, treatment, and prevention services?”* underscores the critical need to address the unique challenges faced by this demographic in accessing SRH services.

Research indicates that migrant and refugee adolescents encounter multifaceted barriers that impede their access to SRH services. A scoping review identified factors such as limited health literacy, cultural stigma, language barriers, and systemic obstacles as significant impediments to HIV prevention and treatment services among migrant youth globally (33). In the Australian context, a participatory action research study involving 87 migrant and refugee youth highlighted socioecological barriers, including lack of awareness about services, cultural disconnects, language barriers, and intergenerational cultural conflicts, as leading to the underutilisation of sexual and reproductive health services (34).

These adolescent-specific access barriers are indicative of a broader pattern affecting SRH service utilisation across the life course. A related concern identified in this study was the question, *“What are the healthcare utilisation patterns, health-seeking behaviours and associated barriers and facilitators (including cultural diversity and beliefs), of this group during pregnancy and the postpartum period?”* This question shifts the focus to maternal care but reflects many of the same structural and cultural obstacles.

Taken together, these findings highlight the need for integrated, culturally responsive SRH strategies that span adolescence through to parenthood. Whether addressing STI prevention or maternal health, equitable access requires the removal of linguistic, cultural, financial, and systemic barriers across all stages of reproductive life. This calls for sustained policy and service-level commitments to inclusivity, continuity of care, and community-informed design.

### Gender-Based Violence Priorities

The prioritisation of the question, *“What are the psychosocial factors (stress, depression, self-esteem, etc*.*) affecting their sexual and reproductive health that also contribute to the perpetration of gender-based violence?”* underscores the complex interplay between mental health and GBV among migrant and refugee populations.

A critical interpretative synthesis study highlights that migrants, particularly forced and irregular migrants, experience high rates of GBV (35). The study identifies unequal power dynamics and fear of disclosure and stigmatisation as key factors exacerbating the psychosocial impact of GBV on migrants. These psychosocial stressors can influence both victimisation and perpetration of GBV within migrant communities. Furthermore, another study examining Arabic-speaking forced migrant women in Melbourne, Australia, reveals that structural and symbolic violence during resettlement significantly impacts mental health and increases the risk of intimate partner violence (IPV) (36). The research emphasises the experiences of structural inequality and culture.

These findings collectively suggest that addressing psychosocial factors such as stress, depression, and self-esteem is crucial in understanding and mitigating GBV among migrant and refugee populations. Interventions must consider the mental health needs of these communities to effectively address and prevent GBV.

The question, *“What are the barriers and facilitators to reporting gender-based violence within this group, and how can healthcare settings identify and respond to this violence?”* highlights the critical challenges faced by migrant and refugee individuals in disclosing GBV incidents. Research findings show that survivors often remain silent due to fear of losing their job, partner or residency status (37). Adolescent girls and other marginalised individuals face heightened risks and fewer options for safe disclosure due to intersecting vulnerabilities (38). Moreover, because many violence-prevention programmes focus exclusively on women or younger children, adolescent girls often fall through the cracks and do not have access to and capital required to advocate for their needs (38). Together, these findings underscore the urgent need for age- and context-specific, survivor-centred approaches within healthcare and community settings that promote safe, confidential, and culturally responsive mechanisms for GBV disclosure and support.

Notably, this question was rated as a high priority by both youth and professional interest-holders, signalling a rare point of convergence and affirming the shared recognition of its critical importance across experiential and institutional domains.

### Perinatal Mental Health and Integrated Care Priorities

The prioritisation of the question, *“Does investing in women’s mental health after pregnancy improve health (physical and mental) outcomes?”* is strongly supported by growing global evidence underscoring the far-reaching impact of perinatal mental health. According to the WHO’s 2022 guide on integrating perinatal mental health into maternal and child health services, up to one in ten women in high-income countries and one in five women in low- and middle-income countries experience mental health conditions during or after pregnancy, with serious implications for both maternal wellbeing and infant development including risks of premature birth, low birth weight, and long-term cognitive and emotional challenges in children (39). The WHO emphasises that early identification and intervention within existing maternal health platforms is not only feasible but essential to breaking this intergenerational cycle of poor health.

Findings from a trial in Japan provide further empirical backing (40). This community-based intervention integrated mental health care into routine maternal and child health services, resulting in significantly improved postpartum mental health outcomes for women in the intervention group compared to those receiving standard care. These results underscore the measurable benefits of investing in mental health during the perinatal period, not only for maternal recovery but for the child’s early development and the broader health system’s effectiveness.

## Data Availability

The data that support the findings of this study are available on request from the corresponding author.

## STRENGTHS AND LIMITATIONS

To the best of our knowledge, this is one of the few exemplars of the PSP approach being applied in Australia (41) and the first to focus specifically on identifying SRH research priorities for migrant and refugee youth populations. Uniquely, the priorities from youth and professional interest-holders were assessed and analysed separately, allowing for a nuanced understanding of differing perspectives. Our approach engaged diverse interest-holders, including young people, clinicians, community representatives, and policymakers, ensuring representative co-production of priorities aligned with end-user needs. The methodological framework employed iterative, transparent, and consensus-driven processes, enhancing result validity and relevance. Most importantly, this study addresses the systematic underrepresentation of marginalised populations in research priority-setting, potentially informing more equitable allocation of research resources and the development of culturally responsive interventions.

Several methodological limitations warrant consideration. First, the contemporary context necessitated predominant reliance on digital data collection methods, potentially limiting engagement from digitally disadvantaged participants. Second, despite comprehensive interest-holders’ recruitment strategies, some subgroups within heterogeneous migrant and refugee communities may remain underrepresented. Third, the Australia-specific focus means findings should be interpreted with caution when considering application to other geopolitical contexts with different healthcare systems, policy frameworks, and migration patterns. These limitations suggest opportunities for complementary priority-setting exercises targeting specific underrepresented subpopulations and international comparative studies to establish cross-contextual validity of identified priorities.

## CONCLUSION/IMPLICATIONS

The 11 research priorities identified through this systematic PSP highlight critical knowledge gaps in SRH specific to migrant and refugee adolescents and young people in Australia. These empirically derived priorities represent a convergence of lived experience and professional insight, illuminating concerns that remain underrepresented in conventional research agendas. The findings underscore a substantive misalignment between existing evidence and the issues prioritised by communities, reinforcing the need for methodologically rigorous and community-engaged research processes. In particular, the observed divergence in how youth and professionals assess SRH needs points to the importance of centring youth perspectives in future agenda-setting.

These priorities offer a clear foundation for the strategic realignment of research investment, policy, and programming toward approaches that are more inclusive, culturally responsive, and equitable. Advancing this agenda will be essential to improving SRH outcomes for marginalised populations in both research and practice. Importantly, the identified priorities provide actionable guidance to researchers and policymakers, enabling targeted study designs and resource allocation that directly address community-identified needs. By integrating these priorities into future research planning, there is potential to foster more effective interventions, inform culturally sensitive policies, and ultimately enhance sexual and reproductive health equity for migrant and refugee youth.

## Acknowledgement

We sincerely thank all the participants, community organizations, and partner institutions who contributed their time and insights to this study. We also acknowledge the valuable guidance and support provided by our steering committee members and collaborators throughout the research process.

## List of abbreviation

GBV: Gender-Based Violence
HIV: Human Immunodeficiency Virus
HREC: Human Research Ethics Committee
ICDP: International Conference on Population and Development
IPV: Intimate Partner Violence
JLA: James Lind Alliance
LGBTQ+: Lesbian, Gay, Bisexual, Transgender, Queer/Questioning, and others
NGO: Non-Governmental Organisation
PMNCH: Partnership for Maternal, Newborn, and Child Health
PSP: Priority Setting Partnership
REPRISE: Reporting guideline for priority setting of health research
SA Health: South Australia Health
SHINE SA: Sexual Health Information Networking and Education South Australia
SRH: Sexual and Reproductive Health
SRHR: Sexual and Reproductive Health and Rights
STD: Sexually Transmitted Disease
WHO: WHO Organization

