## Supplementary file 1 & 2 for "Interest Holders-Driven Research Priorities in Sexual and Reproductive Health for Migrant and Refugee Adolescents and Young Adults in Australia"

**Supplementary file 1: Reporting guideline for priority setting of health research**

| No | REPRISE Item | How It Was Addressed in Our Study |
|---|---|---|
| A | **Context and scope** | |
| 1 | Geographical scope | National-level study focused on migrant and refugee adolescents and young adults in Australia. |
| 2 | Health area, field, focus | Sexual and reproductive health (SRH), covering a broad range of domains including literacy, service access, risk factors, interventions, and settings. |
| 3 | The intended beneficiaries | Adolescents and young adults (aged 18–24) from migrant and refugee backgrounds in Australia. |
| 4 | Target audience of the priorities | Researchers, policymakers, healthcare providers, funders, and community organizations involved in SRH. |
| 5 | Research area | Health services research and psychosocial aspects within SRH |
| 6 | Type of research questions | Etiology, service delivery, health literacy, psychosocial factors, interventions, barriers and facilitators. |
| 7 | Time frame | Interim priorities with potential for future updates; no fixed expiry but intended to guide near- to mid-term research and policy. |
| B | **Governance and team** | |
| 8 | Leadership selection and structure | Steering committee with researchers, clinicians, and young adult representatives overseeing the process. |
| 9 | Characteristics of the team | Multidisciplinary expertise in SRH, migrant/refugee health, adolescent health; includes healthcare providers and youth representatives. |
| 10 | Training or experience relevant to conducting priority setting | Steering committee and team experienced in priority-setting methods and qualitative research, using JLA methodology. |
| C | **Framework for priority setting** | |
| 11 | Framework used | James Lind Alliance (JLA) Priority Setting Partnership methodology. |
| D | **Stakeholders or participants** | |
| 12 | Define the inclusion criteria for stakeholders involved in priority-setting | Youth aged 18–24 from migrant/refugee communities; professionals from academia, healthcare, policy, and NGOs engaged in SRH. |
| 13 | Strategy or method for identifying and engaging stakeholders | Recruitment through partnerships with community organizations, healthcare providers, schools, social media, direct outreach, and steering committee networks. |
| 14 | Number of participants and/or organizations involved | 92 participants in the initial survey: 31 youth interest-holders, 61 professional interest-holders. |
| 15 | Characteristics of stakeholders | Detailed demographics including gender, WHO region of origin, sector affiliation for professionals; youth primarily aged 18–24, diverse cultural backgrounds. |
| 16 | reimbursement for participation | N/A |
| E | **Identification and collection of research priorities** | |
| 17 | Methods for collecting initial priorities | Rapid systematic literature review combined with online surveys presenting 83 evidence uncertainties for rating by participants. |
| 18 | Methods for collating and categorizing priorities | Structured thematic analysis by steering committee, consolidating uncertainties into 12 thematic areas and 5 overarching domains. |
| 19 | Methods and reasons for modifying (removing, adding, reframing) priorities | Steering committee refined, consolidated, and removed duplicates based on scope, clarity, and relevance. |

| | | |
|---|---|---|
| **20** | Methods for refining or translating priorities into research topics or questions | Steering committee reviewed and confirmed research questions after initial survey, prior to final survey and consensus process. |
| **21** | Methods for checking whether research questions or topics have been answered | Rapid literature review assessed current evidence to identify gaps and ensure priorities represent unanswered questions. |
| **22** | Number of research questions or topics | 83 uncertainties initially identified; refined to 18 key priorities after first survey; 11 final priorities confirmed after consensus. |
| **F** | **Prioritization of research topics/questions** | |
| **23** | Methods and Criteria for prioritizing research topics or questions | 9-point Likert scale surveys with median scores calculated separately for youth and professionals; criteria based on perceived importance and consensus. |
| **24** | Method or threshold for excluding research topics/questions | Research questions with low median scores excluded from further rounds; only high/moderate priority questions progressed. |
| **G** | **Output** | |
| **25** | State the approach to formulating the research priorities | Final priorities presented as clear research questions grouped by domain; combined and group-specific priority levels reported. |
| **H** | **Evaluation and feedback** | |
| **26** | Evaluation of prioritization process | Pilot testing of surveys with steering committee and target groups; iterative feedback incorporated to refine instruments and process. |
| **27** | feedback to stakeholders and/or to the public; and how feedback (if received) was addressed and integrated | Summary of priorities and process shared with steering committee; consultation and consensus meetings held |
| **I** | **Implementation** | |
| **28** | Outline the strategy or action plans for implementing priorities | Priorities intended to guide future research, policy, and program development for migrant/refugee youth SRH; engagement with relevant stakeholders for uptake. |
| **29** | plans, strategies, or suggestions to evaluate impact | N/A |
| **J** | **Funding and conflict of interest** | |
| **30** | State sources of funding | 2023 Faculty of Health and Medical Sciences Early Grant Development Award (University of Adelaide Internal Grant Scheme) |
| **31** | Declare any conflicts or competing interests | No conflicts declared in provided text (add statement if relevant). |

**Supplementary file 2**

| **1** | Domain 1: SRH literacy/knowledge | |
|---|---|---|
| | This domain explores the SRH knowledge and understanding that adolescents and young adults from refugee and migrant backgrounds have and the confidence they have in navigating the SRH care system. This domain also aims to understand how SRH literacy (understanding of medical information and how they access and use it to benefit themselves) can be improved amongst this group.<br>All of the questions in the questionnaire refer to **adolescents** and **young adults** (18–24 years) | |
| | **Questions** | **Theme** |
| 1. | How can SRH literacy and access to health information be improved for this group? | Sexual and Reproductive Health needs and services |
| 2 | What are the knowledge gaps and misconceptions regarding family planning and contraception within this group? | FP and contraception |
| 3 | What are their experiences and preferences in terms of contraceptive methods and family planning decision-making? | FP and contraception |
| 4 | What are the perceptions and attitudes of parents and guardians towards family planning and contraception for their children? | FP and contraception |
| 5 | Are there knowledge gaps and misconceptions regarding STIs/STDs/HIV and abortion? | Sexually transmitted infection (STI) or diseases (STD) and HIV |
| 6 | How can health literacy and access to information on pregnancy and maternal health be improved? | Pregnancy and maternal health |
| 7 | What are the perceptions and attitudes of male adolescent's background towards gender equality and gender-based violence prevention in the context of SRH? | Men sexual and reproductive health (SRH) |
| 8 | What are the experiences and outcomes of sexual and reproductive health education and programs for males within this group? | Men sexual and reproductive health (SRH) |
| 9 | What are the knowledge, attitudes, and beliefs of males regarding sexual and reproductive health? | Men sexual and reproductive health (SRH) |
| 10 | How can community-based programs (such as youth mentoring and after-school programs) effectively promote positive sexual and reproductive health outcomes among males? | Men sexual and reproductive health (SRH) |

| 2 | Domain 2: SRH service needs, access and delivery | |
|---|---|---|
| | This domain explores the SRH service access patterns and needs of adolescents and young adults from refugee and migrant backgrounds, including access to counselling, contraception, health care etc. This domain also explores whether the current SRH services are delivered in a comprehensive manner that is culturally sensitive to this unique and vulnerable group. | |
| | **Questions** | **Theme** |
| 1. | What are the major SRH needs, services and preferences of this population group?<br>Access and Navigation to Youth-Friendly SRH Care<br>Family Planning (FP) and Contraception<br>Sexually Transmitted Infection (STI) or Diseases (STD) and HIV<br>Abortion Services and Support<br>Pregnancy and Maternal Health Care<br>Gender-Based Violence Prevention and Support<br>Men's Sexual and Reproductive Health (SRH)<br>Psychosexual, Behavioural, and Mental Health Related to SRH<br>Cultural and Language-Sensitive SRH Services<br>Fertility and Infertility Support<br>Information and Education on SRH<br>LGBTQ+ Specific SRH Services and Support | Sexual and Reproductive Health needs and services |
| 2. | What specific SRH service delivery styles are most impactful and cost effective?<br>Peer-led programs<br>Community-based clinics<br>School-based health services<br>Telehealth services<br>Mobile clinics<br>Youth-friendly health centers<br>Hospital-based services<br>Online platforms and apps<br>Outreach and community education | Sexual and Reproductive Health needs and services |
| 3 | How can access to and use of SRH services within the health system can be improved for this group? | Sexual and Reproductive Health needs and services |
| 4. | What are the experiences and perceptions of adolescents regarding the accessibility and availability of SRH care services and how can community outreach programs and peer support networks enhance service accessibility? | Access and navigation to SRH care |

| | | |
|---|---|---|
| 5. | Is ensuring culturally appropriate sexual health services (e.g. STI/STD/HIV services, including testing, treatment, counselling, and prevention) is necessary? | Sexually transmitted infection (STI) or diseases (STD) and HIV |
| 6. | Is involving community organisations in improving STI/STD/HIV services crucial for this group? | Sexually transmitted infection (STI) or diseases (STD) and HIV |
| 7. | What are the major pregnancy and maternal health needs and challenges faced by this group? | Pregnancy and maternal health |
| 8. | What are their experiences and perceptions of the quality of antenatal/postnatal care services received? | Pregnancy and maternal health |
| 9. | What are the major risk factors for the inequalities and disparities in pregnancy and maternal health outcomes? | Pregnancy and maternal health |
| 10 | What are the best models of care and services in order to ensure culture-sensitive and respectful maternal health and delivery services? | Pregnancy and maternal health |
| 11 | How can quality maternal care (ANC, delivery, PNC) be provided to prevent morbidity and mortality among pregnant/postpartum migrant and refugee adolescent mothers? | Pregnancy and maternal health |
| 12 | Are there specific models of care that works best to improve pregnancy and maternal health outcomes within this group, and what models are proven as most adaptable and cost-effective? | Pregnancy and maternal health |
| 13 | What are the gaps and limitations in existing support services and systems for addressing gender-based violence amongst this group? | Gender based violence |
| 14 | What are the sexual and reproductive health needs and challenges of male adolescents of migrant and refugee background? | Theme: Men sexual and reproductive health (SRH) |
| 15 | What are the healthcare utilisation patterns and experiences of migrant and refugee adolescent males regarding SRH, including HIV and STIs? | Theme: Men's sexual and reproductive health (SRH) |
| 16 | What are the patterns, barriers, and facilitators to accessing culturally sensitive and inclusive sexual and reproductive health services for migrant and refugee adolescent males? | Theme: Men sexual and reproductive health (SRH) |
| 17 | What are the major mental health service needs, barriers, and facilitators to accessing SRH related mental health services (abortion care, sexual violence etc.) and what are their experiences of these services? | Mental Health |
| 18 | What are the major SRH needs and challenges faced by LGBTQ+ adolescents within the group? | SRH of LGBTQ+ |
| 19 | How can SRH care providers address the specific needs and challenges of LGBTQ+ migrant and refugee adolescents? | SRH of LGBTQ+ |

| 3 | Domain 3: SRH risk factors, vulnerability, and outcomes | |
|---|---|---|
| | This domain explores the main risk factors that adolescents and young adults from refugee and migrant backgrounds face in their SRH, including cultural, economic, social and structural challenges. We also explore the SRH outcomes (including STIs/HIV, early pregnancy etc.) of this group and how they are influenced by their unique risk factors. | |
| | **Questions** | **Theme** |
| 1. | What are the SRH risk vulnerabilities experienced by this group? | Sexual and Reproductive Health needs and services |
| 2 | What are the social (employment status, housing etc.), structural (legal status, migration pathway etc.) and cultural determinants that influence their access to SRH care? | Access and navigation to SRH care |
| 3 | What are the key factors, including socio-economic (housing, income etc.), cultural beliefs and social norms, that influence the attitudes, utilisation, and continuation of contraception (including long-acting reversible contraception) use in this group? | FP and contraception |
| 4 | What are the experiences and outcomes of unintended pregnancies among this group, and how can they be prevented through improved family planning and contraception services? | FP and contraception |
| 5 | Is addressing the risk factors and determinants contributing to higher rates of STIs/STDs/HIV among migrant and refugee adolescents in Australia more crucial compared to the general population? | Sexually transmitted infection (STI) or diseases (STD) and HIV |
| 6 | What are the short-term and long-term reproductive health outcomes, psychological well-being, and social implications of abortion? | Abortions |
| 7 | What are the important risk factors associated with abortions among this group? | Abortions |
| 8 | What are the social (employment status, housing etc.) and structural determinants (legal status, migration pathway etc.) that influence their pregnancy and maternal health outcomes? | Pregnancy and maternal health |
| 9 | What are the major determinants of pregnancy complications and maternal health issues and what long-term impacts do they have on their socioeconomic trajectories (education, job prospects etc.)? | Pregnancy and maternal health |
| 10 | What factors are associated with adverse pregnancy and perinatal outcomes (preterm, low birth weight, stillbirths) and how can we predict and prevent these? | Pregnancy and maternal health |
| 11 | What are the risk factors and long-term (mental and physical health) impacts of gender-based violence on adolescents and their families? | Gender-based violence |
| 12 | What are the psychosocial factors (stress, depression, self-esteem etc.) affecting their sexual and reproductive health that also contribute to the perpetration of gender-based violence? | Gender-based violence |
| 13 | What are the psychosocial factors (stress, depression, self-esteem etc.) affecting the sexual and reproductive health of males and what are their experiences regarding reproductive rights and decision making? | Men sexual and reproductive health (SRH) |

| | | |
|---|---|---|
| 14 | What are the long-term reproductive health outcomes for males within this group, including fatherhood experiences and family planning involvement? | Men sexual and reproductive health (SRH) |
| 15 | How does migration status, cultural identity and norms intersect with the sexual and reproductive health experiences and decision-making behaviours of males? | Men's sexual and reproductive health (SRH) |
| 16 | What are the long-term impacts of poor sexual and reproductive health outcomes (e.g., unplanned pregnancy, abortion, STIs) on the mental health of this group? | Mental Health |
| 17 | How are cultural sensitivity and acculturation processes impacting mental health needs and help-seeking behaviours? | Mental Health |
| 18 | What are the barriers and facilitators to accessing culturally sensitive and inclusive SRH care for males? | Culture Sensitive |
| 19 | What are the socio-demographic (age, gender, ethnicity etc.) and sociocultural factors (education, family, etc.) influencing fertility decision-making and what are the implications of these choices? | Fertility and infertility |
| 20 | What are the factors influencing their decision-making regarding fertility preservation options (i.e.. Egg and sperm freezing)? | Fertility and infertility |
| 21 | How does migration status, cultural identity and gender expectations intersect with fertility and infertility experiences? | Fertility and infertility |
| 22 | What are the factors influencing the utilisation of sexual and reproductive health services among LGBTQ+ males? | SRH of LGBTQ+ |

| 4 | Domain 4: SRH interventions (at all levels) | |
|---|---|---|
| | In this domain we aim to explore the strengths, weaknesses and gaps in the different programs (interventions) that exist that aim to improve the SRH of adolescents and young adults from refugee and migrant backgrounds. These interventions could include those mandated by the Government (such as regular vaccinations and health care checks, access to Medicare etc.) or those proposed by local communities or education sectors (health planning services and community meet ups etc.). | |
| 1. | Which SRH programs (such as education, vaccines, health planning, environment changes etc.) are necessary to improve the SRH outcomes in this group and how can they do this in a culturally sensitive manner? | Sexual and Reproductive Health needs and services |
| 2 | What are the effective approaches for conducting SRH research and engaging with this group? | Sexual and Reproductive Health needs and services |
| 3 | How can digital health technologies, such as telehealth and mobile applications, enhance SRH knowledge and accessibility of family planning and contraception? | Access and navigation to SRH care |
| 4 | Is promoting condom use and safe sex practices crucial for protecting them from STIs/STD/HIV? | Sexually transmitted infection (STI) or diseases (STD) and HIV |
| 5 | How could involving families in abortion care reduce stigma for this group? | Abortion |
| 6 | How can community-based programs and peer support networks be utilized to promote positive pregnancy and maternal health outcomes? | Pregnancy and maternal health |
| 7 | How do social support networks and community resources impact the experiences and prevention of gender-based violence? | Gender-based violence |
| 8. | How do girl-centred programs equip adolescent girls to prevent and reduce their risk of gender-based violence? | Gender-based violence |
| 9. | How can adolescent SRH services help to detect, prevent, and mitigate the occurrence of gender-based violence? | Gender-based violence |
| 10. | How do social support networks and community resources impact their experiences of fertility and infertility? | Fertility and infertility |
| 11. | How can community-based programs effectively address their fertility and reproductive health needs? | Fertility and infertility |

| 5 | Domain 5: SRH settings, service use and barriers/challenges. | |
|---|---|---|
| | This domain explores the challenges that adolescents and young adults from migrant and refugee backgrounds have in accessing SRH care and services, including cultural and language barriers as well as legal and migration status barriers. We also explore the reasons why some SRH services work better than others for this group and what needs to be done better to improve service use and appropriate cultural settings of the services. | |
| | **Question** | **Theme** |
| 1. | What are the specific barriers and challenges in accessing SRH care services? | Access and navigation to SRH care |
| 2. | What strategies can help overcome cultural and language barriers and improve healthcare providers' services cultural competence and sensitivity to better meet their SRH care needs? | Access and navigation to SRH care |
| 3. | What are the gender-specific barriers and challenges faced by girls in accessing SRH care services in Australia? | Access and navigation to SRH care |
| 4. | How can the cultural diversity within the migrant and refugee adolescent population be effectively considered and addressed in the planning and delivery of SRH care services? | Access and navigation to SRH care |
| 5. | What are the FP and contraception related barriers and challenges, including migration-related factors such as language proficiency, for this group? | FP and contraception |
| 6. | How can barriers related to cost and accessibility of abortion services be addressed? | FP and contraception |
| 7. | How can community engagement and participation be fostered to develop culturally appropriate family planning and contraception program? | FP and contraception |
| 8. | What are the barriers for migrant and refugee adolescents in understanding and accessing STI/STD/HIV testing, treatment, and prevention services? | Sexually transmitted infection (STI) or diseases (STD) and HIV |
| 9. | How can we better tailor comprehensive SRH education programs to be sensitive to culturally and linguistically diverse adolescents? | Sexually transmitted infection (STI) or diseases (STD) and HIV |
| 10. | What are the barriers and challenges, including migration-related factors such as legal status and cost, faced by girls in accessing safe and legal abortion services, and how can we address these barriers? | Abortion |
| 11. | What are their experiences and perspectives of their autonomy regarding abortion decision-making? | Abortion |
| 12. | What are the healthcare utilisation patterns, health-seeking behaviours and associated barriers and facilitators (including cultural diversity and beliefs), of this group during pregnancy and the postpartum period? | Pregnancy and maternal health |
| 13. | How can healthcare providers promote culturally appropriate and inclusive birthing experiences? | Pregnancy and maternal health |
| 14. | How can early, routine screening for violence in pregnant adolescent women increase disclosure and improve perinatal outcomes? | Pregnancy and maternal health |

| | | |
|---|---|---|
| 15. | What is the magnitude and patterns of gender-based violence among this group, and how do gender norms and cultural expectations contribute to these patterns? | Gender-based violence |
| 16. | What are the barriers and facilitators to reporting gender-based violence within this group, and how can healthcare settings identify and respond to this violence? | Gender-based violence |
| 17. | How does cultural identity and migration status intersect with experiences of gender-based violence among migrant and refugee adolescents? | Gender-based violence |
| 18. | What are the healthcare utilisation patterns and access barriers to sexual and reproductive health services among males? | Men's sexual and reproductive health (SRH) |
| 19. | Does investing on women's mental health through a women-centred approach during and after pregnancy improve health (physical and mental) outcomes? | Mental Health |
| 20. | What are the experiences and perspectives of healthcare providers in delivering culturally sensitive and inclusive SRH care to migrant and refugee adolescents? | Culture Sensitive |
| 21 | What are the specific fertility and infertility challenges faced by this group? | Fertility and infertility |
